# Opportunities and challenges in access to healthcare for international migrants with work-related diseases and injuries in Gulf Cooperation Council countries: A systematic literature review protocol

**DOI:** 10.1101/2025.03.17.25324135

**Authors:** Weijun Yu, Aleena Dawer, Jeanetta Floyd, Nicole Saad, Jiaqin Wu, Katherine O. Robsky, Oliver Johnson, Yulia Hutsul, Dylan Ratnarajah, Bryan Shaw, Martine Etienne-Mesubi, Deus Bazira

## Abstract

**Introduction:** International labor migrants are crucial to the global workforce in the Gulf Cooperation Council (GCC) countries, which host over 11.7% of the world’s migrant workforce, posing significant healthcare challenges. This systematic review aims to evaluate whether international migrants in GCC countries have effective access to healthcare for work-related diseases and injuries and to propose evidence-based recommendations for policy and healthcare interventions.

**Methods:** This review will include studies from 2013 to 2023 published in peer-reviewed journals in English or Arabic (with English abstracts) available on PubMed, Embase and CINAHL. We focus on healthcare access for work-related diseases and injuries among international migrants in GCC countries. Search strategies are developed using MeSH terms and key terms related to our study population (international immigrants), context (the GCC countries), and exposure (migrant status; work-related diseases and injuries). The screening process involves two stages: initial review of titles/abstracts and full-text review. Studies meeting eligibility criteria and focus on our outcome (access to healthcare) will be included. Data extraction will cover study characteristics, population demographics, described exposures, outcomes measured, and key findings. Given the expected heterogeneity, we will primarily use a narrative synthesis approach, with meta-analysis as an optional method.

**Discussion:** Our systematic review aims to assess how GCC countries manage healthcare access for international migrants. By considering both migrant workers and expatriate professionals, we provide a culturally tailored perspective. Methodological rigor is ensured through the gold standard screening process, where at least two reviewers independently screen the literature at each stage, with a senior reviewer resolving discrepancies. Our review will identify barriers and facilitators, informing targeted interventions for policymakers. Ultimately, our findings will support evidence-based strategies to improve healthcare access for international migrants’ in GCC countries.

**Systematic review registration:** This systematic review protocol was registered on PROSPERO (CRD42024532851) on April 21, 2024.

## Introduction

International labor migrants, encompassing both international migrant workers and expatriate professionals, form a critical segment of the global workforce. International migrant workers are individuals aged 15 and over, who relocate from their country of origin to participate in the labor force of their host country, significantly contributing to economies, particularly in regions like the Arab States [1]. Expatriate professionals, on the other hand, are highly skilled individuals employed for their specialized skills, abilities, and experience, engaging in global careers [2], [3]. Regardless of their essential roles, the health necessities of International labor migrants are frequently overlooked [4].

International migrant workers often undertake low-wages and high-risk jobs, exposing them to hazardous conditions; over 150 million migrant workers globally face conditions that lead to elevated prevalence of occupational morbidity and mortality [5], [6].International labor migrants encounter significant barriers to healthcare access in their host countries including limited or no health insurance, restricted rights to statutory health care [7], [8], language barriers, and unfamiliarity with local healthcare infrastructure [9]. A previous systematic review and meta-analysis study covering Asia, the Middle East, Europe and the Americas shows that globally, about 47% of international migrant workers suffer from occupational health issues, and 22% report workplace injuries or accidents [10]. The International Labour Organization (ILO) reports that approximately 7600 people die daily from work-related injuries or illnesses, with 15% of deaths directly attributable to work-related injuries [11]. Despite calls from International organizations like the World Health Organization for governments and policymakers worldwide to adopt strategies that enhance occupational health for International labor migrants, the implementation of such strategies remains limited [7], [12–14].

The Gulf Cooperation Council [15] countries (Bahrain, Kuwait, Oman, Qatar, Saudi Arabia, and the United Arab Emirates) constitute the largest international hub for migrant labor, hosting nearly 30 million migrants [16], which accounts for more than 11.7% of the global migrant workforce [5]. Among these countries, Qatar, Kuwait, and the United Arab Emirates (UAE) have the highest percentage of migrants [17], [16]. This significant influx of labor migrants present substantial healthcare challenges, exacerbated by the transient nature of migration in the region. For instance, the protection of international workforce rights and safety in Qatar is rooted in the kafala system, a sponsorship system that requires employers to protect the rights of foreign employees [18]. Despite its original good intention, the system is controversially associated with labor control and exploitation, which may have led to poor health outcomes, including the mortality of 6,500 migrant workers during the 2022 FIFA World Cup [18].

Labor-intense workers are predominantly employed in high-risk jobs such as construction and agriculture, where they are exposed to extreme heat, leading to severe health issues such as cardiac arrest and exhaustion [19], [20]. A significant number of these migrant workers coming from Southeast Asia, with India alone experienced nearly 34,000 deaths of Indian migrant workers in the GCC countries from 2014 to 2019 due to such conditions [21]. The Covid-19 pandemic further underscored the vulnerability of these labor migrants, shifting the risk from merely physical injuries to now infectious diseases [22], [7]. Concurrently, the rapid economic development in the GCC countries has led to an increase in work-related injuries among the expatriate professionals, contributing significantly to non-fatal injuries [23]. While each GCC country has its own healthcare regulations concerning labor force, the pandemic exacerbated the situation, leaving many individuals vulnerable, without access to essential medical service, or unable to return home for care [24–26].

From a patient-centered perspective, access to healthcare is defined as the ability to identify, seek, reach, obtain, and utilize healthcare services, ensuring that patients’ needs are met [27]. This concept integrates multiple dimensions of service acquisition and delivery, including availability, appropriateness, preference, and timeliness, to provide effective care for patients [28]. Achieving satisfactory access to healthcare is characterized by several factors: having health insurance that simplifies integration into the healthcare system, being able to seek necessary services promptly, or having a consistent healthcare provider to build a strong patient-provider relationship [29].

Given the critical role that international migrants play in the economic development of the GCC countries, it is crucial to address the healthcare opportunities and challenges they face. At the time of writing, no systematic review has investigated whether international migrants experiencing work-related diseases and injuries in the GCC countries have efficient and effective access to healthcare services. Our study aims to bridge this gap, identify the barriers and facilitators impacting their access, and propose evidence-based recommendations to inform policies and healthcare practice, ultimately enhancing health outcomes for one of the most essential yet vulnerable populations within the GCC countries.

### Objectives

The objective of this systematic review is to comprehensively summarize findings on whether international migrants experiencing work-related diseases and injuries in the GCC countries have efficient and effective access to healthcare services. Our goal is to identify and propose evidence-based recommendations that will inform the development of policies and healthcare interventions aimed at enhancing access to healthcare services for international migrants facing work-related diseases and injuries in the GCC countries.

### Research questions

Our research question is structured following the Population, Exposure, Outcome (PEO) framework. We consider international migrants in the GCC countries as our population, with migrant status and their experience of work-related diseases and injuries as our exposures, and access to healthcare services as our outcome. Our main research question is “Whether international migrants experiencing work-related diseases and injuries in the GCC countries have access to healthcare services, and what factors influence this access over the past decade?”

This review will also cover the following subtopics:

a. Barriers and facilitators impacting timely access to healthcare services
b. Specific policies developed and implemented to address barriers that constrain international migrants’ access to healthcare services
c. Coverage of healthcare services available to international migrants
d. Affordability and/or financing
e. Healthcare service utilization
f. Health workforce compatibility (e.g., language barriers, cultural competence, availability of international migrant-friendly services)
g. Effectiveness (e.g., quality of healthcare service, responsiveness)

## Methods

### Study design

We developed this systematic review protocol in accordance with the Preferred Reporting Items for Systematic Reviews and Meta-Analyses Protocols (PRISMA-P) Checklist [30]. On April 21st, 2024, our protocol was successfully registered on the International Prospective Register of Systematic Review (PROSPERO) with the registration number CRD42024532851 [31]. We will present the full systematic review findings by following the Preferred Reporting Items for Systematic Reviews and Meta-Analyses (PRISMA) 2020 guidelines [32]. A PRISMA flow diagram will be created to describe our process of identification, screening, eligibility, and inclusion of final articles in this systematic review. This study is determined not to involve human subjects, thus ethical review and approval are formally waived by Georgetown University’s Institutional Review Board committee (IRB ID: STUDY00007743).

### Eligibility criteria

To ensure a focused analysis in our systematic review, we developed eligibility criteria outlined in Table 1. We will include peer-reviewed articles written in English or Arabic (with English abstracts available) published between January 1st, 2013 and December 31st, 2023. Our focus will be on access to healthcare for work-related diseases and injuries or occupational health among international migrants in the Gulf Cooperation Council countries (Bahrain, Kuwait, Oman, Qatar, Saudi Arabia, and the United Arab Emirates).

**Table 1:** Eligibility Criteria.

| **Criteria** | **Inclusion** | **Exclusion** |
| --- | --- | --- |
| **Population** | -International migrant workers moved to GCC (Gulf Cooperation Council) countries (Bahrain, Kuwait, Oman, Qatar, Saudi Arabia, and the United Arab Emirates) for labor-intensive employment opportunities, driven by economic necessity.<br>- Expatriate professionals relocate for specialized or managerial roles with competitive benefits. | - International migrants living outside GCC countries.<br>- Local migrants with GCC citizenship status. |
| **Language** | -Literature written in English.<br>-Literature written in Arabic with English abstracts available. | Literature written in other languages without English abstracts. |
| **Time Frame** | Studies conducted between January 1st, 2013 and December 31st, 2023. | Studies conducted before January 1st, 2013. |
| **Study design** | - Primary studies (RCT) providing original research findings in peer-reviewed journals.<br>- Observational studies (cross-sectional, cohort, | Non-peer-reviewed articles, theses, dissertations, book reviews, news articles, |
|  | case-control) in peer-reviewed journals.<br>- Review articles (systematic reviews, meta-analyses, narrative reviews) in peer-reviewed journals.<br>- Editorials, opinion pieces | conference reviews, or reports. |
| **Subject** | Studies evaluating or discussing access to healthcare for work related diseases and injuries. | Studies without specific reference to access to healthcare for work related diseases and injuries. |
| **Accessibility** | Studies or reports where full text is available. | Studies or reports where no full text is available. |

### Information sources

We will search the following three databases for eligible peer-reviewed literature: PubMed, EMBASE, and CINAHL. Full versions of articles meeting our eligibility criteria, based on their titles, abstracts, and key descriptors, will be obtained. Additionally, we will explore references and bibliographies of review articles to identify further relevant articles that may have been missed during the initial screening. RefWorks software will be used to manage references and remove duplicates.

### Search strategy

Search terms, keywords, Medical Subjects Headings (MeSH) terms, and Boolean operators will be used to develop search strategy for each database. We formulated our search algorithm based on terms related to our study population (international immigrants), context (the GCC countries), and exposure (migrant status; work-related diseases and injuries), framed by our research question. Following previous recommendations [33], we did not include our study outcome (access to healthcare) terms in our search strategy to optimize the search results.

### Screening

The screening process will be performed in two stages: 1) an initial review of titles and abstracts, followed by 2) examination of full texts. Five authors from our research team will conduct the screening. Four authors will be divided into two groups, with two reviewers assigned to each stage for each stage. Screening will be independently conducted to identify articles that meet our eligibility criteria. A senior reviewer will serve as the third reviewer to resolve discrepancies and crosscheck results between the two independent reviewers for each stage.

Inter-rater reliability tests will be conducted separately for each stage. Initially, two reviewers will independently assess a sample subset of 20% of studies to align on evaluation criteria and ensure a consistent inter-rater reliability rate of at least 85%. Multiple rounds of inter-rater reliability tests will be held until the expected 85% rate is achieved. After this alignment, the two reviewers will proceed to independently evaluate the remaining articles. Disagreements will be resolved by the senior reviewer to maintain objectivity in study selection. Reasons for excluding each article will be clearly recorded throughout the screening process. The selection process and decisions will be recorded in an Excel spreadsheet for detailed data extraction and analysis. At the time of writing, the literature search for this systematic review has not yet been completed.

### Data extraction

For each included study, our data extraction will cover five domains:

a. Study characteristics (author, year, country, language, study design, sample size),
b. Population demographics (age, gender, origin nationality),
c. Exposures described (Migrant status, work-related disease or injury),
d. Outcomes measured (access to healthcare, timeliness, barriers, facilitators)
e. Key findings (effect sizes, thematic insights).

At least two reviewers will extract data to ensure the accuracy and reliability of the information collected. Discrepancies in data extraction between reviewers will be resolved through discussion, or by involving the third reviewer if necessary to achieve consensus. For missing data or unclear details, we will attempt to contact study authors for clarification or additional information. If authors cannot be reached, we will record missing data as “Not applicable”. Extracted data will be organized and managed using an Excel spreadsheet.

### Risk of bias and quality assessment

In the article appraisal phase, we will utilize the Mixed Methods Appraisal Tool (MMAT) [34] to assess the quality of qualitative, quantitative, and mixed methods research. Each included study will be evaluated for the suitability of its design in addressing our research question, the adequacy of the sample size, participants recruitment bias, outcome measurement accuracy, and the clarity of result reporting. This meta-narrative approach allows for a systematic interpretation and integration of findings across diverse study designs.

For the risk of bias in randomized controlled trials (RCTs), we will apply the Cochrane Collaboration’s Risk of Bias Tool, [35] assessing random sequence generation, allocation concealment, and blinding of participants and personnel. For observational studies, the ROBINS-I (Risk Of Bias In Non-randomized Studies of Interventions) tool [36] will be used to assess biases related to confounding, participants selection, and intervention classification.

### Data synthesis

Considering the expected heterogeneity among our included studies, we will primarily employ a narrative synthesis approach. This approach is chosen due to the anticipated diversity in study designs (qualitative, quantitative, mixed-methods), exposures (migrant status, work-related diseases and injuries), and outcomes (access to healthcare access, barriers, facilitators). Data will be synthesized when at least two studies report similar outcomes, ensuring our conclusions are drawn from robust evidence. Consistency across studies will be assessed based on the direction of effects and the contexts of interventions to determine the reliability of synthesized findings.

We will synthesize findings related to primary outcomes, including healthcare access levels, barriers and facilitators to access. We will systematically categorize and compare studies, identifying common themes and differences. While our primary focus will be on narrative synthesis, if we identify more than two quantitative studies that meet criteria for homogeneity, we will consider conducting a meta-analysis to further quantify the effects and outcomes reported. Statistical methods such as meta-regression may be utilized to explore potential sources of heterogeneity and to provide a better understanding of the data. Additionally, we may employ sensitivity analysis to test the robustness of our findings, ensuring that our synthesis reflects the most reliable evidence available.

## Discussion

Our proposed systematic review has the potential to significantly influence how the GCC countries address access to healthcare for international migrants. One of our strengths is its comprehensive inclusion of the study population. We not only investigate migrant workers but also include expatriate professionals, providing an inclusive and culturally tailored perspective for the GCC countries. Our methodological rigor is enhanced by applying the gold standard of screening, where at least two reviewers independently screen the literature in parallel, rather than splitting it between reviewers to accelerate the process. This approach strengthens the robustness and reliability of our data collection. By recognizing and analyzing specific barriers and facilitators, our review will equip policymakers with the insights needed to promote more targeted interventions to improve timely access to healthcare among international migrants. Additionally, our review will report work-related injuries across various sectors, which is vital for developing equitable healthcare policies that cater to all international migrants, regardless of their job type or industry. As a result, our findings can inform the development of evidence-based policies and healthcare interventions that are specifically designed to enhance access to healthcare services for international migrants in the GCC countries.

Our review investigates the complexities surrounding access to healthcare for international migrants in the GCC countries who experience work-related diseases and injuries. By comprehensively evaluating the literature, we aim to uncover whether these migrants have effective and efficient access to healthcare services and identify the factors influencing this access over the past decade. Our analysis will cover several critical dimensions, including the barriers and facilitators impacting the timeliness of healthcare access, specific policies developed to address these barriers, and the coverage of healthcare services available to international migrants. We will also examine affordability and financing of healthcare services, utilization patterns, and the compatibility of the health workforce, such as language barriers, cultural competence, and availability of migrant-friendly services. Additionally, we will assess the overall effectiveness of healthcare services provided to international migrants, considering the quality of care and system responsiveness.

While our review is designed to be comprehensive and rigorous, there are some limitations that should be acknowledged. The variability in healthcare systems and migrant policies across the GCC countries presents a challenge in standardizing data collection and analysis. However, we will address this by employing robust data synthesis techniques and possibly sensitivity analysis to ensure our findings are as reliable as possible. Another potential limitation is the availability of data. Some information on migrant health may not be publicly available due to political sensitivities. To mitigate this, we will rely on peer-reviewed literature to ensure the credibility of our data. Moreover, our inclusion of studies published in both English and Arabic, along with efforts to contact study authors for missing data, will help to minimize the impact of data availability issues.

Our final systematic review findings will be submitted to high-impact peer-reviewed journals for publication to ensure broad visibility. We will also present our findings at international seminars, conferences, and stakeholders’ meetings to reach a wide audience and stimulate further discussion within professional communities.

## Acknowledgement

We extend our gratitude to Ms. Shannon Mulligan at Georgetown University Center for Global Health Practice and Impact, for her coordination and recruitment of collaborative authors from multidisciplinary backgrounds, which greatly enriched this project.

## Data availability statement

Data will be made available upon the completion of the full systematic review.

## Funding

The first and senior authors are sponsored by Georgetown University Center for Global Health Practice and Impact for their research work compensation. The second author is sponsored by Georgetown University Global Health Institute Student Fellowship program. One collaborative author is sponsored by Georgetown University IDOL Family Summer Fellowship program.

## Competing interests

The authors have declared that no competing interests exist.

## Notes

### Competing Interest Statement

The authors have declared no competing interest.

## References

1. International Labour Organization. ILO Global Estimates on International Migrant Workers Results and Methodology Executive Summary (Third Edition); 2021. Available from: https://www.ilo.org/global/topics/labour-migration/publications/WCMS_808935/lang--en/index.htm.

2. Siljanen T, Lämsä AM. The changing nature of expatriation: Exploring cross-cultural adaptation through narrativity. The International Journal of Human Resource Management. 2009 Jul 1;20(7):1468–86.

3. McNulty Y, Vance CM. Dynamic global careers: A new conceptualization of expatriate career paths. Personnel Review. 2017 Mar 6;46(2):205–21.

4. Abbas M, Aloudat T, Bartolomei J, Carballo M, Durieux-Paillard S, Gabus L, et al. Migrant and refugee populations: a public health and policy perspective on a continuing global crisis. Antimicrobial Resistance & Infection Control. 2018 Dec;7:1–1.

5. Sweileh WM. Global output of research on the health of international migrant workers from 2000 to 2017. Globalization and health. 2018 Dec;14:1–2.

6. World migration report 2018 [Internet]. International Organization for Migration; 2017. Available from https://www.iom.int/wmr/world-migration-report-2018.

7. Aktas E, Bergbom B, Godderis L, Kreshpaj B, Marinov M, Mates D, et al. Migrant workers occupational health research: an OMEGA-NET working group position paper. International Archives of Occupational and Environmental Health. 2022 May 1:1–3.

8. Mladovsky P, Rechel B, Ingleby D, McKee M. Responding to diversity: an exploratory study of migrant health policies in Europe. Health policy. 2012 Apr 1;105(1):1–9.

9. Almutairi KM. Culture and language differences as a barrier to provision of quality care by the health workforce in Saudi Arabia. Saudi medical journal. 2015;36(4):425.

10. Hargreaves S, Rustage K, Nellums LB, McAlpine A, Pocock N, Devakumar D, et al. Occupational health outcomes among international migrant workers: a systematic review and meta-analysis. The Lancet Global Health. 2019 Jul 1;7(7):e872–82.

11. Mehmood A, Maung Z, Consunji RJ, El-Menyar A, Peralta R, Al-Thani H, et al. Work related injuries in Qatar: a framework for prevention and control. Journal of occupational medicine and toxicology. 2018 Dec;13:1–0.

12. Zimmerman C, Kiss L, Hossain M. Migration and health: a framework for 21st century policy-making. PLoS medicine. 2011 May 24;8(5):e1001034.

13. Ahonen EQ, Benavides FG, Benach J. Immigrant populations, work and health—a systematic literature review. Scandinavian journal of work, environment & health. 2007 Apr 1:96–104.

14. Ahonen EQ. Occupational health challenges for immigrant workers. In Oxford Research Encyclopedia of Global Public Health 2019 Oct 30.

15. Secretariat General of the Gulf Cooperation Council. Member States [Internet]. [cited 2024 June 5]. Available from: https://gcc-sg.org/en-us/AboutGCC/MemberStates/Pages/Home.aspx.

16. Rahman MM, Umar S, Awad Almarri S. Healthcare Provisions for Migrant Workers in Qatar. Health & Social Care in the Community. 2023;2023(1):6623948.

17. Sönmez S, Apostopoulos Y, Tran D, Rentrope S. Human rights and health disparities for migrant workers in the UAE. Health and Human Rights. 2013 Aug;2(13):30–68.

18. Alfarizi MH, Morgan KR, Lago MC. Human Rights Abused in Qatar: FIFA Puts World Cup More Than Lives?. Jurnal Penegakan Hukum dan Keadilan. 2023 Sep 30;4(2):112–22.

19. Bener A. Health status and working condition of migrant workers: Major public health problems. International journal of preventive medicine. 2017 Jan 1;8(1):68.

20. Migrant Workers Face Heightened Risk of Death and Injury: New IOM Report. 2021. International Organization for Migration (IOM). Available from: https://publications.iom.int/books/occupational-fatalities-among-international-migrant-workers.

21. Asi YM. Migrant workers’ health and COVID-19 in GCC countries. Arab Center Washington DC. 2020 Jul 7;7.

22. Asiri SM, Kamel S, Assiri AM, Almeshal AS. The epidemiology of work-related injuries in Saudi Arabia between 2016 and 2021. Cureus. 2023 Mar;15(3).

23. Consunji RJ, Mehmood A, Hirani N, El-Menyar A, Abeid A, Hyder AA, et al. Occupational safety and work-related injury control efforts in Qatar: lessons learned from a rapidly developing economy. International journal of environmental research and public health. 2020 Sep;17(18):6906.

24. Rahman MM. COVID-19 and Migrants in the GCC states: Challenges, responses and key lessons.

25. Jureidini R. Global governance and labour migration in the GCC. Global governance and Muslim organizations. 2019:339–64.

26. Nahari TH, Alkhidir MA, Ibrahim HM, Al Mamun M. Migrant workers with COVID-19: a major challenge for Gulf Cooperation Council (GCC) countries to curb the spread of infection. Global Health Promotion. 2024 Jan 5:17579759231216108.

27. Levesque JF, Harris MF, Russell G. Patient-centred access to health care: conceptualising access at the interface of health systems and populations. International journal for equity in health. 2013 Dec;12:1–9.

28. Berry LL, Seiders K, Wilder SS. Innovations in access to care: a patient-centered approach. Annals of internal medicine. 2003 Oct 7;139(7):568–74.

29. 2021 National Healthcare Quality and Disparities Report [Internet]. Rockville (MD): Agency for Healthcare Research and Quality (US); 2021 Dec. Available from: https://www.ncbi.nlm.nih.gov/books/NBK578529/

30. Moher D, Shamseer L, Clarke M, Ghersi D, Liberati A, Petticrew M, et al. Preferred reporting items for systematic review and meta-analysis protocols (PRISMA-P) 2015 statement. Systematic reviews. 2015 Dec;4:1–9.

31. Weijun Yu, Deus Bazira, Aleena Dawer, Jeanetta Floyd, Jiaqin Wu, Katherine Robsky, et al. Opportunities and Challenges in Timely Access to Healthcare for International Migrants with Occupational Diseases and Injuries in Gulf Cooperation Council Countries (Bahrain, Kuwait, Oman, Qatar, Saudi Arabia, and the United Arab Emirates): A Systematic Literature Review. PROSPERO 2024 CRD42024532851 Available from: https://www.crd.york.ac.uk/prospero/display_record.php?ID=CRD42024532851

32. Page MJ, McKenzie JE, Bossuyt PM, Boutron I, Hoffmann TC, Mulrow CD, et al. Updating guidance for reporting systematic reviews: development of the PRISMA 2020 statement. Journal of clinical epidemiology. 2021 Jun 1;134:103–12.

33. Tawfik GM, Dila KA, Mohamed MY, Tam DN, Kien ND, Ahmed AM, et al. A step by step guide for conducting a systematic review and meta-analysis with simulation data. Tropical medicine and health. 2019 Dec;47:1–9.

34. Hong QN, Fàbregues S, Bartlett G, Boardman F, Cargo M, Dagenais P, et al. The Mixed Methods Appraisal Tool (MMAT) version 2018 for information professionals and researchers. Education for information. 2018 Jan 1;34(4):285–91.

35. Higgins JP, Altman DG, Gøtzsche PC, Jüni P, Moher D, Oxman AD, et al. The Cochrane Collaboration’s tool for assessing risk of bias in randomised trials. Bmj. 2011 Oct 18;343.

36. Sterne JA, Hernán MA, Reeves BC, Savović J, Berkman ND, Viswanathan M, et al. ROBINS-I: a tool for assessing risk of bias in non-randomised studies of interventions. bmj. 2016 Oct 12;355.

